# Trunk control status in children with neuromuscular disorders and typically developing children: is there a measurable difference?

**DOI:** 10.1101/2025.03.12.25322083

**Authors:** Tania E. Sakanaka, Penelope B. Butler, Richa Kulshrestha, Tracey Willis, Ian D. Loram

**Author notes:** Correspondence, Address: Department of Life Sciences, Faculty of Science and Engineering, Manchester Metropolitan University; The Dalton Building, Oxford Road, Manchester, M1 5GD, UK.

## Abstract

**Aim:** To test sensitivity of the Segmental Assessment of Trunk Control (SATCo) to changes in neutral vertical (NV) head and trunk control in children with neuromuscular disorders (NMD) and compared with typically developing (TD) children.

**Methods:** In this observational cross-sectional study, SATCo was applied in 19 children with NMD (10y6m(3y7m), 2 female) and 19 TD children (7y10m(4y12m), 6 female). Statistical differences between *condition* (NMD and TD), *segment* (seven trunk segments), and *control type* (static, active, and reactive control), and relationship between age and SATCo outcomes were analysed with linear mixed-effects models (LME).

**Results:** SATCo scores were measurably different between *conditions* (F_(1,39.6)_=151.0,p<.001), *segments* (F_(6,138.3)_=23.3,p<.001), *control type* (F_(2,33.0)_=6.3,p=.005), and interactions *condition segment* (F_(6,138.3)_=23.3,p<.001) and *condition control type* (F_(2,33.0)_=6.3,p=.005). A significant relationship between decreasing active control and increasing age in 10 children with Duchenne muscular dystrophy confirmed measurable decline in trunk control following diagnosis.

**Interpretation:** The results showed that SATCo is sensitive to differences in trunk control status in NMD. While still ambulatory, children with NMD presented already a measurable trunk control deficit at varying segments. SATCo has potential as an outcome measure for therapeutic interventions or effectiveness of treatments promoting NV trunk control in children with NMD and needs further study.

**What this paper adds:**

- Neutral vertical head/trunk control was different between NMD and TD children
- SATCo is sensitive to head/trunk control differences between NMD and TD children
- Ambulant children with NMD showed measurable head/trunk control deficit at various segments
- SATCo detected subtle head/trunk control changes in early disease stages of NMD
- Active segmental trunk control declines with increasing age in Duchenne muscular dystrophy

## Introduction

Neuromuscular disorders (NMDs) encompass a heterogeneous group of conditions affecting different aspects of the motor unit. With onset during childhood, paediatric NMDs are mostly genetic in aetiology and vary in distribution of affected muscles, age of onset, and severity of progression. Disability can be mild to severe depending on how muscle weakness, joint contractures, altered sensory perception, respiratory deficiency, skeletal deformities, impaired ambulation, fatigue, and/or exercise intolerance affect an individual’s function.

Historically, the prognosis for treatment of individuals with these disorders was non-existent. However, this scenario has been changing since the implementation of the standard of care^1–4^ and the introduction of glucocorticoids to slow disease progression in Duchenne muscular dystrophy (DMD)^5^. More recently, disease-modifying therapies for the treatment of spinal muscular atrophy (SMA), among others in development,^6^ are dramatically improving life expectancy and the performance of everyday life activities.

Various functional assessments have been developed for clinical use and, more recently, these have been used as outcome measures from drug therapies.^7^ As the range of conditions and symptoms vary significantly, some scales assess a wide range of activities in ambulant and non-ambulant patients,^8–11^ while others are specific to a subset of disorders and relate to specific development of weakness and contractures.^12–14^

This knowledge is vital for patient management and provides clear, practical information, e.g. the patient remains able to transfer from sitting on a chair to standing. However, these tests are not intended to detect the possible underlying mechanisms of subtle changes in motor ability that occur through all stages of all types of NMDs, especially in the early stages when symptoms are mild or the later stages when patients are non-ambulant and have increased muscle weakness.

Effective head and trunk control is critical to upper and lower limb function and fundamental to ambulation^15–17^ but is not generally a specific assessment in NMD although trunk weakness is a common factor. The Segmental Assessment of Trunk Control (SATCo) is a validated clinical test ^18^ that is sensitive to subtle changes in trunk control status of alignment with the gravitational vertical (neutral vertical posture, NVP).^19^ It tests the head/trunk control in a systematic form by: (i) segmenting to control of six head/trunk segments and full trunk, and (ii) differentiating to *static, active* and *reactive* control. SATCo has been applied previously in DMD and shown to detect changes in trunk control.^20–22^ Importantly, SATCo identifies the topmost (most cephalo) segment at which control of the fully upright NVP is not demonstrated for each type of control. SATCo may thus have potential to provide additional information about head and trunk control status in children with NMD.

The objective of this study was to determine, using SATCo, if there was a measurable difference in trunk control status in children with various types of NMDs and typically developing children (TD). Significant differences between groups would mean that SATCo could potentially be used as a guideline for therapeutic interventions or as a measure of treatment effectiveness on NV trunk control.

## Method

### Study design, setting, and participants

This was an observational cross-sectional study that formed a subset of a larger study (MRC-funded project “Quantification of head and trunk control for children with neuromotor and neuromuscular disorders”, MR/T002034/1).

This convenience subsample of 19 children with a clinical diagnosis of NMD and 19 TD children were screened for eligibility and recruited between 2021-2024 by the clinical staff from four hospitals and by advertising to staff from a university to test their children (TD) (Tables 1 and 2).

**Table 1.**
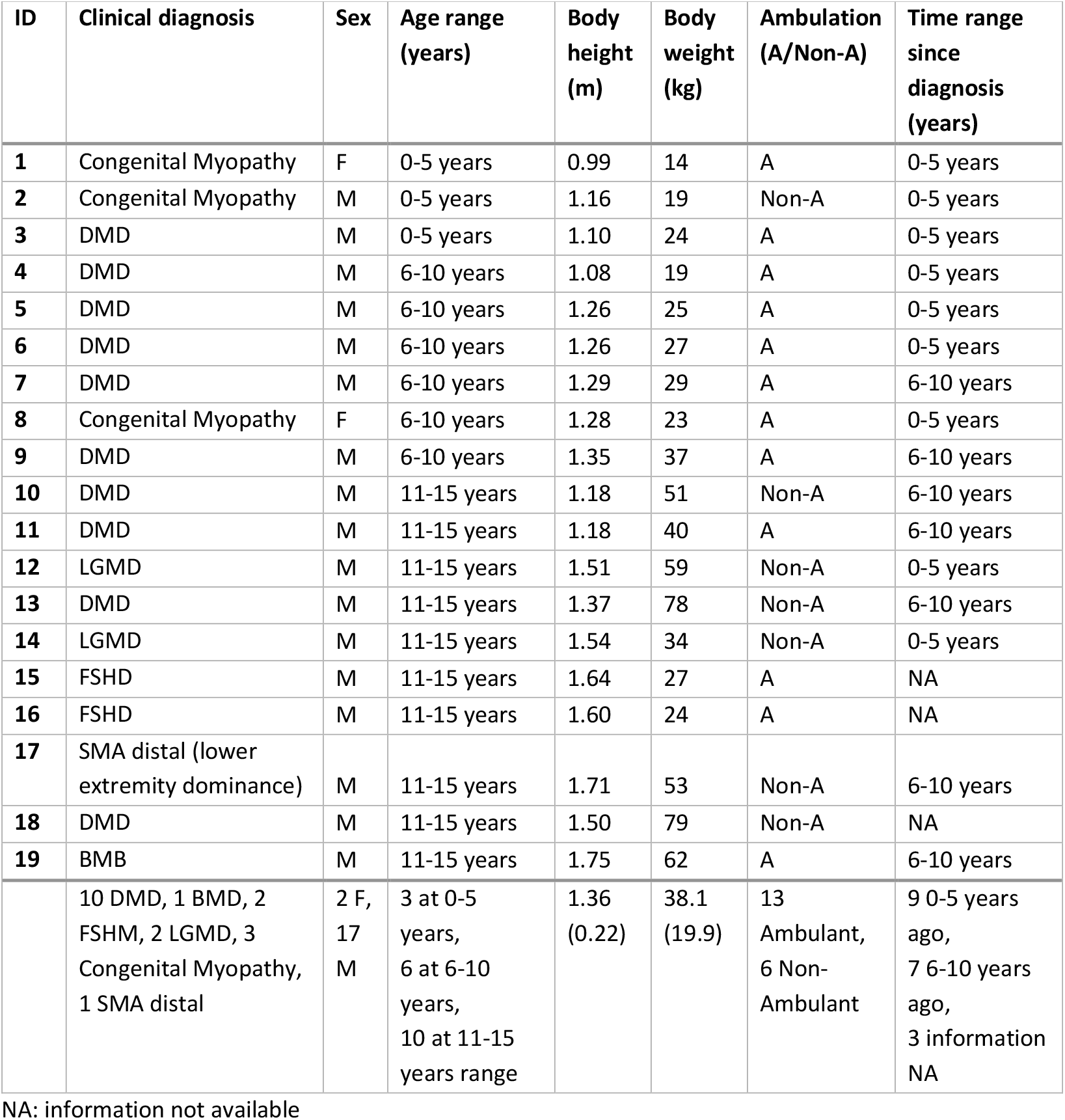
Clinical and anthropometric characteristics of participants with NMD. Age and time since diagnosis are displayed as age-range: 0-5 years, 6-10 years, and 11-15 years.

| ID | Clinical diagnosis | Sex | Age range (years) | Body height (m) | Body weight (kg) | Ambulation (A/Non-A) | Time range since diagnosis (years) |
| --- | --- | --- | --- | --- | --- | --- | --- |
| 1 | Congenital Myopathy | F | 0-5 years | 0.99 | 14 | A | 0-5 years |
| 2 | Congenital Myopathy | M | 0-5 years | 1.16 | 19 | Non-A | 0-5 years |
| 3 | DMD | M | 0-5 years | 1.10 | 24 | A | 0-5 years |
| 4 | DMD | M | 6-10 years | 1.08 | 19 | A | 0-5 years |
| 5 | DMD | M | 6-10 years | 1.26 | 25 | A | 0-5 years |
| 6 | DMD | M | 6-10 years | 1.26 | 27 | A | 0-5 years |
| 7 | DMD | M | 6-10 years | 1.29 | 29 | A | 6-10 years |
| 8 | Congenital Myopathy | F | 6-10 years | 1.28 | 23 | A | 0-5 years |
| 9 | DMD | M | 6-10 years | 1.35 | 37 | A | 6-10 years |
| 10 | DMD | M | 11-15 years | 1.18 | 51 | Non-A | 6-10 years |
| 11 | DMD | M | 11-15 years | 1.18 | 40 | A | 6-10 years |
| 12 | LGMD | M | 11-15 years | 1.51 | 59 | Non-A | 0-5 years |
| 13 | DMD | M | 11-15 years | 1.37 | 78 | Non-A | 6-10 years |
| 14 | LGMD | M | 11-15 years | 1.54 | 34 | Non-A | 0-5 years |
| 15 | FSHD | M | 11-15 years | 1.64 | 27 | A | NA |
| 16 | FSHD | M | 11-15 years | 1.60 | 24 | A | NA |
| 17 | SMA distal (lower extremity dominance) | M | 11-15 years | 1.71 | 53 | Non-A | 6-10 years |
| 18 | DMD | M | 11-15 years | 1.50 | 79 | Non-A | NA |
| 19 | BMB | M | 11-15 years | 1.75 | 62 | A | 6-10 years |
|  | 10 DMD, 1 BMD, 2 FSHM, 2 LGMD, 3 Congenital Myopathy, 1 SMA distal | 2 F, 17 M | 3 at 0-5 years, 6 at 6-10 years, 10 at 11-15 years range | 1.36 (0.22) | 38.1 (19.9) | 13 Ambulant, 6 Non-Ambulant | 9 0-5 years ago, 7 6-10 years ago, 3 information NA |

NA: information not available

**Table 2.** Anthropometric characteristics of TD participants. Age is displayed as age-range: 0-5 years, 6-10 years, 11-15 years, and 16 years.

| ID | Sex | Age range (years) | Body height (m) | Body weight (kg) |
| --- | --- | --- | --- | --- |
| 1 | M | 0-5 years | 0.82 | 13 |
| 2 | M | 0-5 years | 0.89 | 14 |
| 3 | M | 0-5 years | 1.00 | 15 |
| 4 | M | 0-5 years | 1.03 | 16 |
| 5 | F | 0-5 years | 1.09 | 19 |
| 6 | M | 0-5 years | 1.14 | 18 |
| 7 | M | 0-5 years | 1.08 | 18 |
| 8 | M | 0-5 years | 1.16 | 21 |
| 9 | M | 6-10 years | 1.23 | 26 |
| 10 | M | 6-10 years | 1.27 | 26 |
| 11 | F | 6-10 years | 1.29 | 32 |
| 12 | M | 6-10 years | 1.34 | 32 |
| 13 | F | 6-10 years | 1.38 | 29 |
| 14 | M | 6-10 years | 1.36 | 28 |
| 15 | M | 6-10 years | 1.29 | 26 |
| 16 | F | 11-15 years | 1.51 | 42 |
| 17 | F | 11-15 years | 1.59 | 42 |
| 18 | F | 11-15 years | 1.68 | 60 |
| 19 | M | 16 years | 1.76 | 59 |
|  | 6 F,<br>13 M | 8 at 0-5 years,<br>7 at 6-10 years,<br>3 at 11-15 years,<br>1 at 16 years | 1.26 (0.25) | 28.2(13.9) |

Inclusion criteria were children with NMD and TD children between 2 and 16 years old; exclusion criteria were surgical fixation of the spine, both parents or guardians with insufficient understanding of English to give consent, and children who posed a physical risk during testing to themselves or to the research team.

Information about the study and opportunity for questions to be discussed/answered were given before parents/guardians signed a written informed consent. Ethical approval was granted by the London-Brent Research Ethics Committee (IRAS project ID: 233469).

SATCo, body height and weight, and time since diagnosis were recorded.

### Experimental protocol

SATCo was conducted by two trained assessors. The child was seated on a Leckey Therapy Bench (James Leckey Design Ltd, Lisburn, Ireland) adjusted to ensure hip and knee flexion at 90 degrees and the pelvis stabilised in neutral and safely secured to the bench with the provided pelvic support.

The first assessor provided firm horizontal manual support to the trunk, sequentially lowering this support to test each of the seven segments: H (*head*) – support at the shoulder girdle, UT (*upper-thoracic*) - axillae, MT (*mid-thoracic*) - inferior scapula, LT (*lower-thoracic*) - over lower ribs, UL (*upper-lumbar*) - below ribs, LL (*lower-lumbar*) - pelvis, and FT (*full trunk*) - no support given.

Three forms of postural control were tested for each segment:

1. *Static*: maintenance of unsupported NVP for 5 seconds
2. *Active*: maintenance of NVP during a child-initiated head turn to left and right.
3. *Reactive*: the ability to return to NVP following a nudge from the front, back, and both sides, performed by the second assessor (Not tested at the *head* segment).

Control was defined as Demonstrated, Not Demonstrated, or Not Tested for each segment and each type of control.

Demonstration of control was confirmed if the child was able to maintain the trunk segments above the manual support in NVP without external support. Such external support included upper limb support e.g. arms touching their own body or the bench or external support from another person. The inability to maintain NVP was scored as Not Demonstrated.

Control was considered Not Tested if there was an assessor error, for example, assessor preventing child aligning or assessor not holding segment steady in space. Attainment of NVP is a pre-requisite for the *reactive control* test.

SATCo^23^ conventionally ceases testing once the status of each form of control is established. For this exploratory study, testing continued to reveal any more caudal variances in NVP control status.

Three Intel® RealSense™ Depth Cameras D435 were used with camera positions adjusted to record the participant’s lateral right (20Hz), diagonal front left (20Hz), and diagonal back right views (6Hz) (Fig.1).

**Figure 1.**
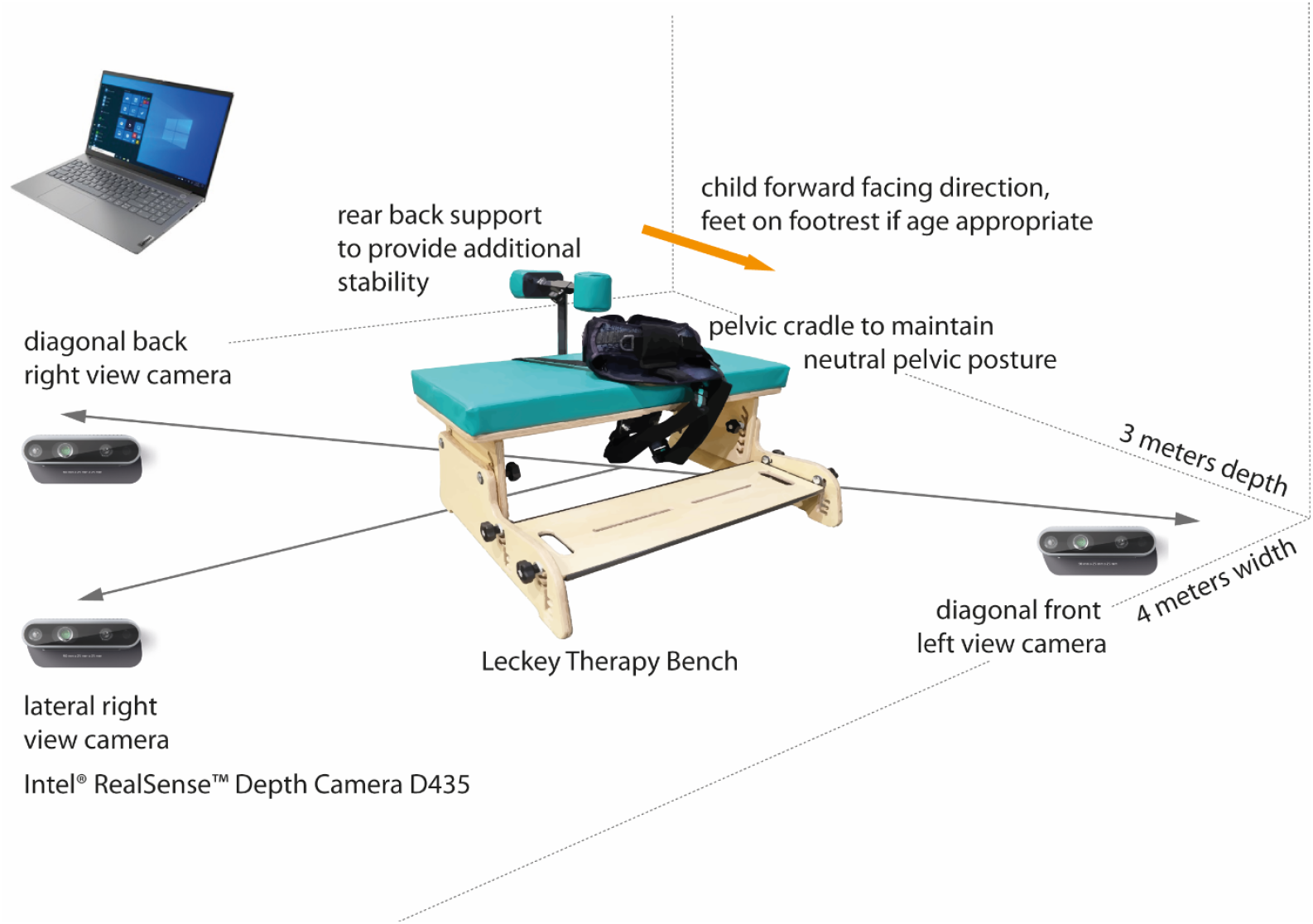
Experimental setup

The child’s SATCo performance was scored according to published criteria^23^ by PBB and TES, authors, and TP, a contributor to this study, from the recorded videos.

### Statistical analysis

The software IBM® SPSS® Statistics for Windows version 28.0.1.1, 2022 (IBM Corp., Armonk, NY, USA), was used for statistical analysis. Age differences were tested with a linear mixed effects model (LME) and sex differences were tested with the Chi-square test.

Statistical tests report, at alpha=0.05, the outcome of a SATCo assessment (Control Demonstrated, Control Not Demonstrated, Not Tested) using LME: 38 children (19 NMD, 19 TD) with factors *condition* (NMD, TD), *segment* (Head, UT, MT, LT, UL, LL, FT), *control type* (static, active, reactive), included within fixed and random effects (grouped by subject), and ANOVA (Satterthwaite approximation) using the SPSS function ‘MIXED’. SATCo outcomes were also reported with *age* as covariate in children with DMD only.

## Results

Nineteen children with NMD and 19 TD children participated in this study (Tables 1 and 2). No significant differences were found between NMD and TD groups in sex, *X*^2^_(1,38)_ =2.53, p=.11. Average age was lower in TD, F_(1,36)_=4.39, p<.05. This would not affect the comparison between groups, as most TD children achieve full NV trunk control at 12 months.^24^

Three main groups of NMDs were tested: muscular dystrophies (ten children with DMD, two with facioscapulohumeral muscular dystrophy – FSHD, two with limb–girdle muscular dystrophy – LGMD, and one with Becker muscular dystrophy – BMD), myopathies (three children with congenital myopathy), and motor neuron disorders (one child with distal spinal muscular atrophy – SMA). Age at diagnosis varied depending on the type of NMD (Table1).

Thirteen out of 19 children with NMD (68.4%) demonstrated NV control in at least one segment and condition, and control was not demonstrated in at least one segment and condition in all 19 children, regardless of clinical diagnosis and ambulation status (Fig.2).

**Figure 2.**
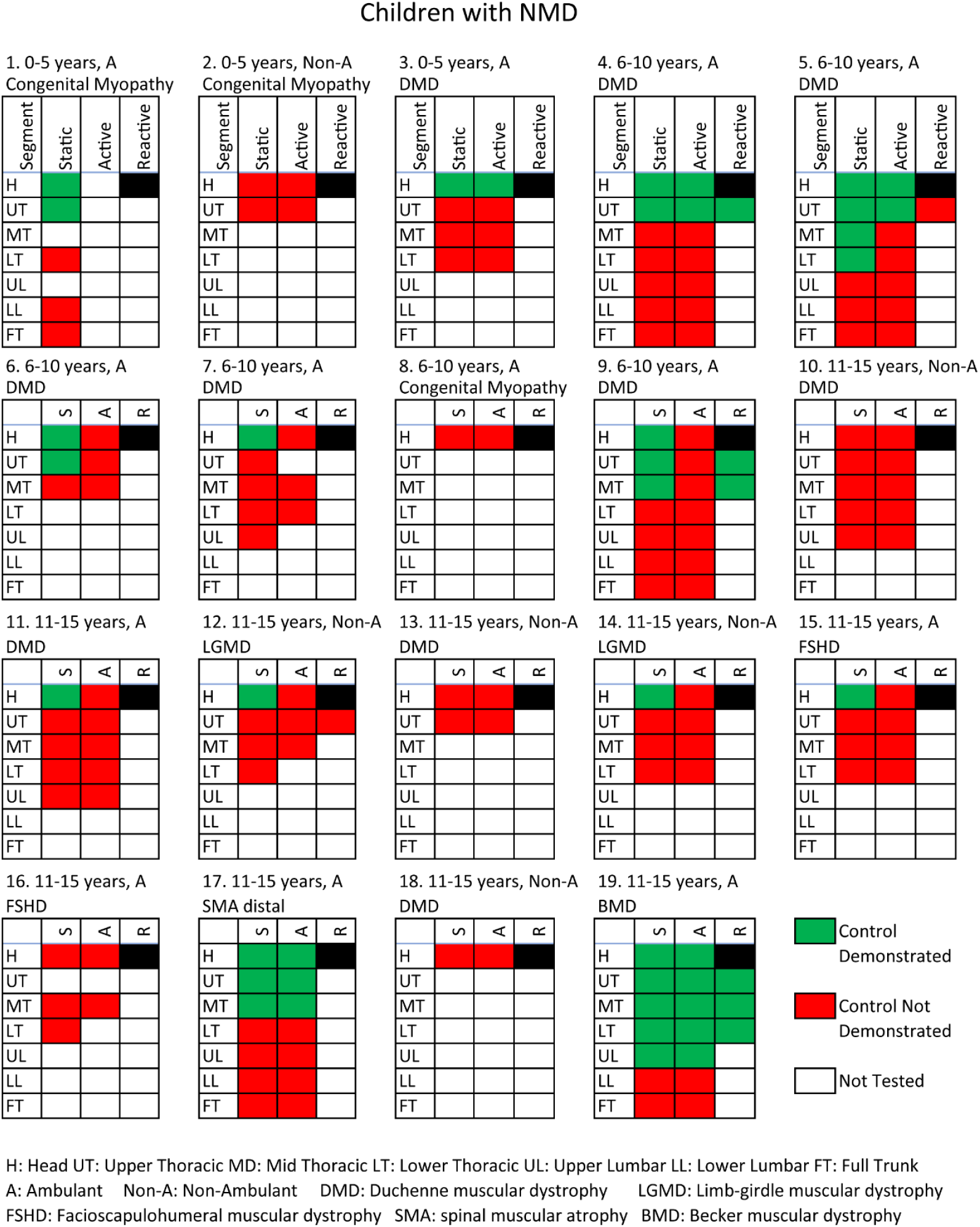
SATCo scores from participants with NMD. Participants are organised by age range, starting from top-left.

Thirteen of the 19 children with NMD (68.4%) were ambulant but even these children presented a measurable trunk control deficit. An ambulant child with congenital myopathy and another with FSHD demonstrated no NVP control at any head/trunk segment. The other 11 ambulant children demonstrated control, but only at the higher trunk segments. Among them, seven children with DMD, one with SMA distal, one with FSHD, and one with congenital myopathy demonstrated NV control only at head or at head and thoracic segments (Fig.2). In general, static control was demonstrated more often, then active, followed by reactive control. Finally, only the child with BMD, a milder form of NMD, demonstrated NV control from the head through the upper lumbar segment.

In the group of non-ambulant children, only two with LGMD demonstrated NVP head static control. The remaining, three with DMD and one with congenital myopathy, did not demonstrate NVP control at any trunk segment.

Interestingly, all seven ambulant children with DMD demonstrated at least one segment of NVP postural control, while none of the three non-ambulant children with DMD demonstrated control at any segment (Fig.3). A significant relationship between increased age and decreased active control was found in DMD (F_(1,8)_=9.3,p<.05). Static control results were leaning towards significance (F_(1,8)_=9.1,p=.08).

**Figure 3.**
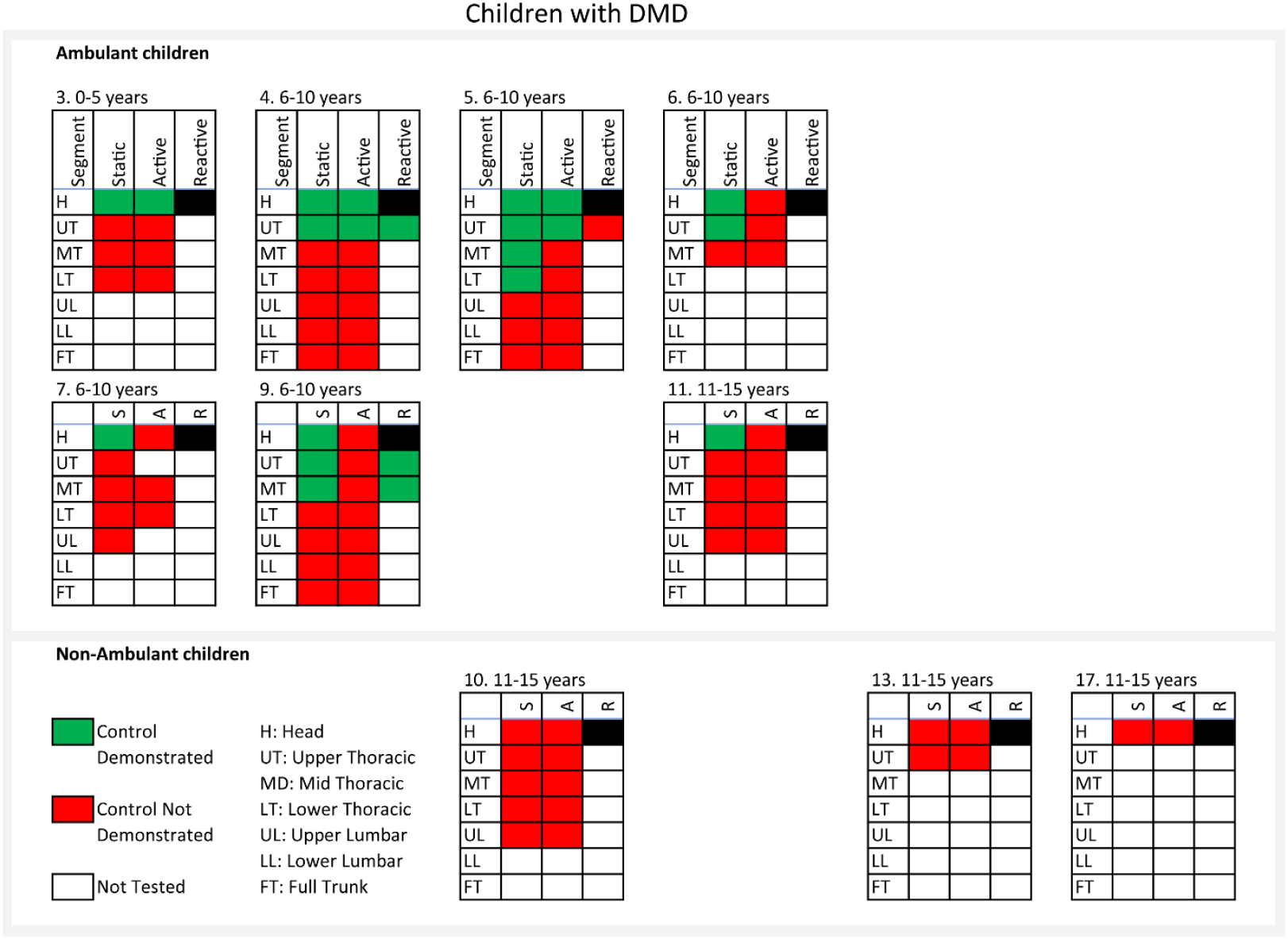
SATCo scores from participants with DMD only. Participants are organised by age range, starting from top-left.

All TD children were able to demonstrate full NVP trunk control (Fig.4).

**Figure 4.**
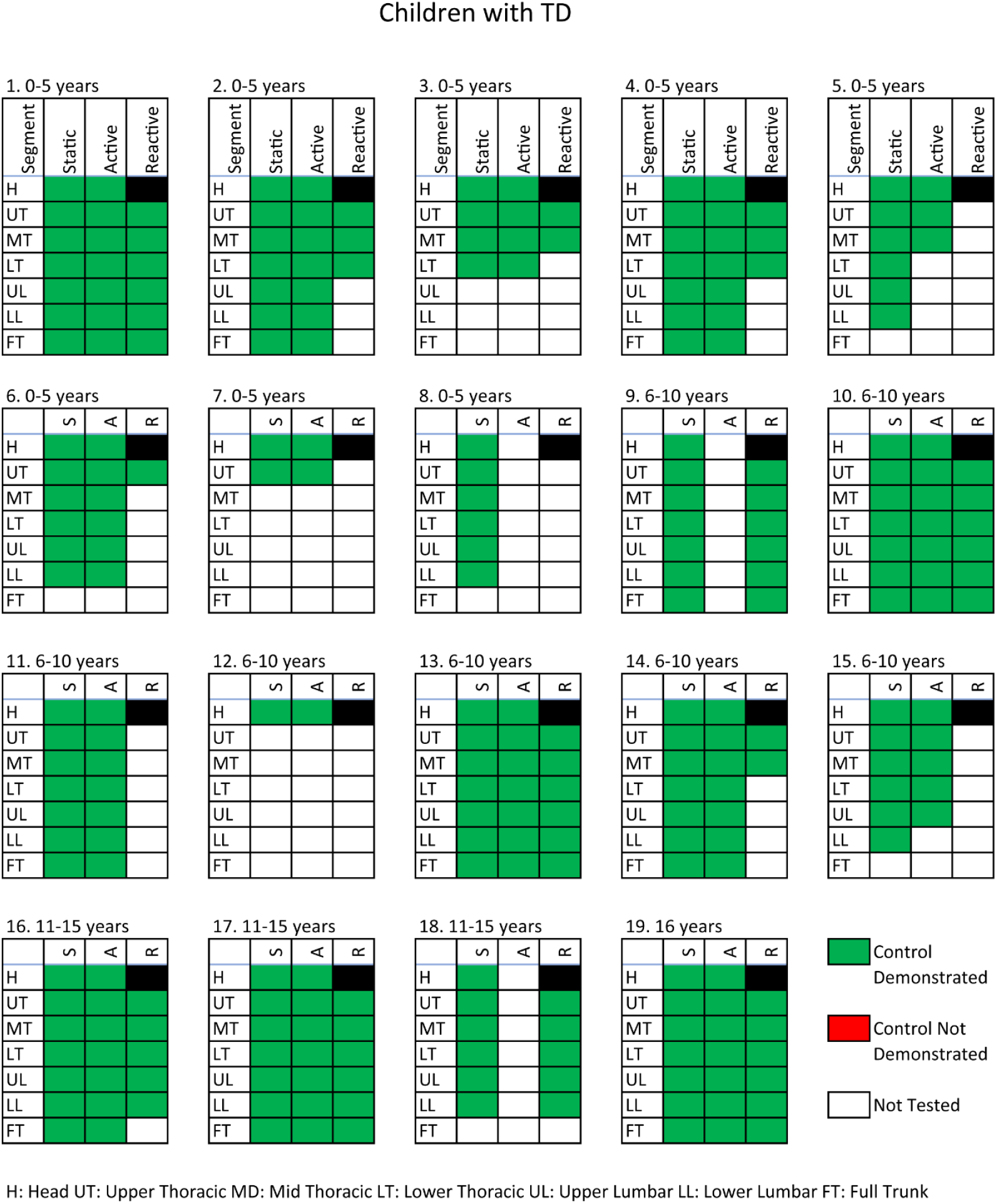
SATCo scores from TD participants. Participants are organised by age range, starting from top-left.

The results from LME analysis confirm that SATCo scores from NMD participants were measurably different from the ones recorded from TD participants (F_(1,39.6)_=151.0,p<.001). Effects of *segment* (F_(6,138.3)_=23.3,p<.001), and *control type* (F_(2,33.0)_=6.3,p=.005), interactions *condition* x *segment* (F_(6,138.3)_=23.3,p<.001) and *condition* x *control type* (F_(2,33.0)_=6.3,p=.005) were also significant. This confirms that differences in SATCo scores between NMD and TD children were related to segment and type of control being tested (Fig.5).

**Figure 5.**
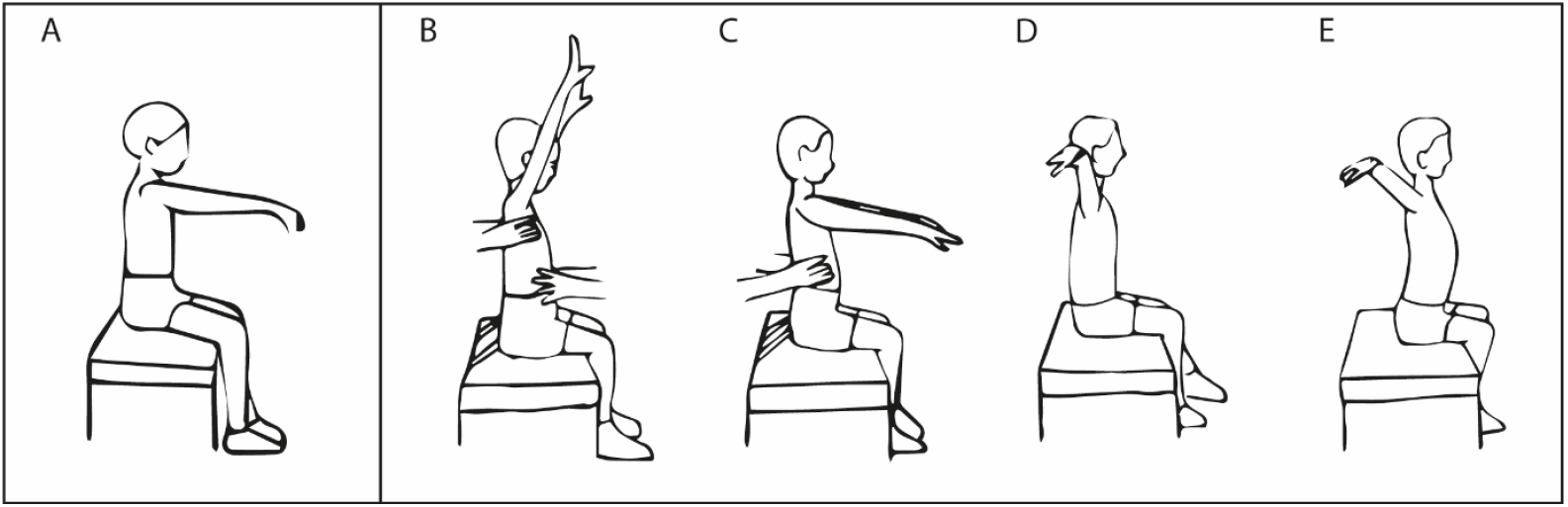
Examples of testing procedures in SATCo. All examples were created from tracing images of recorded data. **A**. Full trunk static control demonstrated by a TD participant. Figures B, C, D, and E were created from recordings of the same ambulant DMD participant (ID #3). **B**. Upper thoracic static control demonstrated. **C**. Lower thoracic static control not demonstrated, participant leaning backward. **D**. Full trunk static control not demonstrated, slight flexion of the lower thoracic and lumbar spine. **E**. Full trunk static control not demonstrated, extension of the thoracic and lumbar spine. The participant was not able to achieve and maintain a neutral vertical posture (see reference image A) but switched between these two extremes (D and E). Thus, although he clearly has sitting ability, his trunk control difficulties are evident.

## Discussion

This study compares NVP segmental trunk control in 19 children with NMD and 19 age-matched TD children. A significant difference in SATCo outcomes between groups, and a significant difference in results from each segment and type of control was found.

### Explanation about SATCo Not Tested results

This data is a subset of a larger dataset being used to train a neural network to automate SATCo outcomes and provide feedback to assessors regarding the interpretation of SATCo. The present study is setting a rigorous standard for the objective SATCo which will be encoded in the neural network enabling future clarity of outcomes.

The incidence of Not Tested scores reflects the rigour of this assessment. SATCo assesses a child’s NVP independent head/trunk control above the manual support, i.e. with the upper limbs and head free of external support. These criteria ensure that head/trunk control alone is tested and not with support provided, for example, by the upper limbs. Due to the strict nature of SATCo, there will be inevitably some instances of Not Tested segments due to tester error. This reflects the reality of assessing children, especially young lively TD children, and the assessor’s skill in performing SATCo (Fig.4).

### SATCo detects trunk control deficit while children with NMD are ambulant

SATCo tests the child’s ability to regulate the position of unsupported head/trunk segments so they remain in or close to NVP by active muscular effort in all directions of movement available at the relevant joint(s). If follows that control is Not Demonstrated if achieved by the use of internal passive support i.e. taking a joint to end of range/maintaining the position through ligament tension or use of external passive support from the upper limbs or contact with external objects.^19^

It is noteworthy that the results for children with NMD showed a measurable trunk control deficit at varying segments: this confirms that there is some degree of control deficit in the trunk while children with NMD are still ambulant (Fig.5). This may be enhanced by weakness of relevant muscles. This deficit in trunk control might explain some of the compensatory strategies that children with NMD use while performing functional activities.^17^ In walking, for example, many children with NMD present an extreme lumbar lordosis: this strategy would take the lumbar joints to their end of range of motion, so providing a ‘bony/ligament end-stop’ meaning that a stable trunk posture can be maintained with minimal muscle activity.

Previous studies also demonstrated deficits in trunk control in ambulant and non-ambulant children with DMD when tested with SATCo.^20–22^ The results from these studies confirm a gradual trend from higher to lower ability to control a sequential number of trunk segments from ambulant to non-ambulant DMD. However, these results differ from our results with respect to trunk control ability. None of the ambulant children with DMD (n=7) in our study demonstrated full trunk control, whereas in Santos et al.,^22^ 65.5% (n=19) demonstrated full trunk control. This discrepancy might be due to our smaller sample size, and/or a higher rigour of applying SATCo in our study. It is noted that Santos and co-authors found a high incidence of scoliosis in their patient groups but it is not known if this was a fixed bony change or ‘postural’ i.e. passively correctable. It is possible that lack of clarity around SATCo testing in this situation may have contributed to their high number of non-ambulant patients showing full trunk control.

### Importance of using functional assessments as a measure of treatment effectiveness in NMD

For individuals with NMD, it is natural to assume that outcomes related to motor function are the most useful measure of drug effect. Ultimately, the objective of any treatment is to maintain or improve function and minimise disability if cure is not possible. The use of the North Star Ambulatory Assessment (NSAA)^14^ as a measure of drug effectiveness in DMD is justifiable, as it was designed specifically to assess the progressive loss of the ability to walk in boys with DMD, the main debilitating characteristic of the disorder.^25^

Conway et al.,^26^ applied the framework provided by the International Classification of Functioning, Disability and Health (ICF) from the World Health Organization (WHO) to identify a core set of items from ICF most relevant for the assessment of function in DMD and BMD. In a group of 703 individuals, in items related to neuromusculoskeletal and movement-related functions, 66% presented limitations in joint mobility (KAFO/AFO/night splint, tendon release), 99% in muscle power (Gowers’ sign, inability to keep up with peers, independent ambulation ceased, muscle hypotonia, muscle weakness, transfer assistance, trouble climbing stairs, trouble walking, wheelchair use), and 86% in gait pattern (abnormal gait, toe walking).

Translating a plethora of motor measures into a single short assessment that can be used in the clinical setting is a challenge. SATCo takes a different approach by evaluating the underlying mechanisms of trunk control that are essential for effective function, including upper and lower limb function.^15–17^ Furthermore, it can be applied to individuals across all stages of any type of NMD, with no prerequisites such as sitting ability, thus including both ambulant and non-ambulant phases.^22^

A SATCo assessment might take about 10-30 min to be completed depending on child ability.

### SATCo is sensitive to differences in NVP trunk control between NMD and TD not detected in other functional assessments

Functional assessments commonly used in the clinical setting are not sensitive to the specific subtle differences in NV head/trunk control detected by SATCo, particularly important to test during the short window between diagnosis and loss of ambulation when there is more opportunity to alter disease progression. A correlation between increased age and decreased active trunk control was found in children with DMD, confirming that SATCo shows how much trunk control worsens as disease progresses.

Among the most recommended assessments^27,28^, the Motor Function Measure (MFM)^11^ scores sitting control on a chair (item 9). This item treats the trunk as a single unit and does not consider the possibility that the component trunk segments may show different control statuses. It corresponds with the highest level of ability tested by SATCo, full trunk control.

The NSAA does not include items that specifically assess NV head and trunk control. The PUL2 will give an indication of trunk control but includes components that have arm/hand support e.g. hands lap to table. This confounds the assessment of trunk control in isolation. The Brooke^8^ and Vignos^9^ Scales broadly rank function. The *Egen Klassifikation* (EK)^10,29^ scale is specific for non-ambulatory individuals and independent sitting is not scored.

## Limitations

This was a small exploratory study, hence the small sample size. Nevertheless, the results clearly indicate a marked difference between children with NMD and TD children.

## Conclusion and future studies

The results from this study support the proposal that SATCo is sensitive to differences in trunk control between NMD and TD children. A correlation between decreased active control and increased age found in DMD confirmed that trunk control worsens as disease progresses, showing that SATCo detects disease progression.

This work also raises initial questions about what changes occur in trunk control in NMD following diagnosis and when these changes commence. A greater understanding of disease progression could assist in a more objective identification of appropriate interventions and their timing and duration during the initial stages of the disease, when there is greatest opportunity to alter progression of the condition.

Although SATCo requires some moderate training of staff who implement the test, its value in terms of resulting information should ensure its place in future research. Linking SATCo findings to the outcomes of functional tests has great promise. There may also be the potential for the use of SATCo during clinical appointments to evaluate drug therapy. The larger study (MR/T002034/1) of which the data reported here is a subset, is producing a clinically based neural network to provide objective analysis and feedback regarding its implementation. This objective SATCo (oSATCo) should facilitate training and delivery of SATCo assessment.

Future studies, including longitudinal studies and randomized clinical trials, are recommended to map the progressive segmental changes in trunk control that occur post-diagnosis in NMD and to facilitate a greater understanding of intervention effects.

## Data Availability

The data that support the findings of this study will be available to data scientists using the images for medical imaging analysis research. The decision on whether to supply will reside with Prof Ian Loram and the review panel from the MRC-funded project MR/T002034/1. The criterion to supply will require the users to demonstrate an information security policy equivalent to this project. Appropriate licenses together with data availability statements will outline these conditions and terms of use, as well as access protocols.

## Abbreviations

BMD: Becker muscular dystrophy
DMD: Duchenne muscular dystrophy
FSHD: Facioscapulohumeral muscular dystrophy
H, UT, MT, LT, UL, LL, FT: Head, Upper thoracic, Mid thoracic, Lower thoracic, Upper lumbar, Lower lumbar, Full trunk
LGMD: Limb–girdle muscular dystrophy
LME: Linear mixed-effects model
SATCo, oSATCo: Segmental Assessment of Trunk Control, Objective Segmental Assessment of Trunk Control
SMA: Spinal muscular atrophy
MD: Muscular dystrophy
NMD: Neuromuscular disorder
NSAA: North Star Ambulatory Assessment
NV, NVP: Neutral vertical, neutral vertical posture
TD: Typically developing

## Acknowledgments

We gratefully acknowledge Teresa Perez (TP) and Natalia Rajkowska from Manchester Metropolitan University for their contributions to this study. We also thank James Leckey Design Ltd (Lisburn, Ireland) for kindly providing Leckey Therapy Benches for this study.

